# Estimating hepatitis B virus cccDNA persistence in chronic infection

**DOI:** 10.1101/2020.02.04.20020362

**Authors:** Katrina A. Lythgoe, Sheila F. Lumley, Lorenzo Pellis, Jane A. McKeating, Philippa C. Matthews

**Affiliations:** Big Data Institute, University of Oxford, Old Road Campus, Oxford, OX3 7LF; Department of Zoology, University of Oxford, Medawar Building, South Parks Road, Oxford OX1 3SY; Nuffield Department of Medicine, University of Oxford, Medawar Building, South Parks Road, Oxford OX1 3SY; Department of Infectious Diseases and Microbiology, Oxford University Hospitals NHS Foundation Trust, John Radcliffe Hospital, Headley Way, Oxford OX3 9DU; Department of Mathematics, University of Manchester, Manchester, UK; Nuffield Department of Medicine Research Building, University of Oxford, Oxford, OX3 7LF; NIHR Biomedical Research Centre, John Radcliffe Hospital, Headley Way, Oxford OX3 9DU

**Keywords:** hepatitis B virus, cccDNA, persistence, evolution, dynamics, reservoir, modelling

## Abstract

Hepatitis B virus (HBV) infection is a major global health problem with over 240 million infected individuals at risk of developing progressive liver disease and hepatocellular carcinoma. HBV is an enveloped DNA virus that establishes its genome as an episomal, covalently closed circular DNA (cccDNA) in the nucleus of infected hepatocytes. Currently available standard-of-care treatments for chronic hepatitis B (CHB) include nucleos(t)ide analogues (NA) that suppress HBV replication but do not target the cccDNA and hence rarely cure infection. There is considerable interest in determining the lifespan of cccDNA molecules to design and evaluate new curative treatments. We took a novel approach to this problem by developing a new mathematical framework to model changes in evolutionary rates during infection which, combined with previously determined within-host evolutionary rates of HBV, we used to determine the lifespan of cccDNA. We estimate that during HBe-antigen positive (HBeAg^POS^) infection the cccDNA lifespan is 61 (36-236) days, whereas during the HBeAg^NEG^ phase of infection it is only 26 (16-81) days. We found that cccDNA replicative capacity declined by an order of magnitude between HBeAg^POS^ and HBeAg^NEG^ phases of infection. Our estimated lifespan of cccDNA is too short to explain the long durations of chronic infection observed in patients on NA treatment, suggesting that either a sub-population of long-lived hepatocytes harbouring cccDNA molecules persists during therapy, or that NA therapy does not suppress all viral replication. These results provide a greater understanding of the biology of the cccDNA reservoir and can aid the development of new curative therapeutic strategies for treating CHB.

**Author Summary:** Nearly one million people die each year due to hepatitis B virus (HBV) related diseases. Although antiviral treatments for HBV exist, cure is rare and treatment is typically life-long, reflecting the persistence of episomal copies of the viral DNA (cccDNA) in the liver. Our knowledge of the cccDNA reservoir in chronic hepatitis B (CHB) is limited. HBV has a high mutation rate and the key determinants of cccDNA dynamics can be inferred by examining the rate of viral evolution. Combining a mathematical model and known rates of HBV evolution we estimate the cccDNA lifespan during different phases of CHB. Our results provide important insights into the dynamics of the HBV reservoir that will inform the design of future treatment interventions.

## INTRODUCTION

Hepatitis B Virus (HBV) is a global health problem, with more than 240 million chronically infected individuals at risk of developing liver fibrosis, cirrhosis, and hepatocellular carcinoma [1]. HBV is the prototypic member of the *hepadnaviruses*, a family of small, enveloped hepatotropic viruses with a partially double-stranded relaxed circular DNA (rcDNA) genome that replicates via reverse transcription of pregenomic RNA (pgRNA). Following the infection of hepatocytes, the rcDNA genome is imported to the nucleus and converted to covalently closed circular DNA (cccDNA), that provides the transcriptional template for pregenomic and subgenomic RNAs. Newly synthesized pgRNA is assembled into nucleocapsids that undergo reverse transcription to generate rcDNA, which is subsequently enveloped and released as infectious virions. Alternatively, capsids can be redirected to the nucleus to replenish and maintain the episomal pool of cccDNA and this intracellular amplification pathway, together with the long half-life of cccDNA, contributes to viral persistence [2,3].

Chronic hepatitis B (CHB) is usually treated with nucleos(t)ide analogues (NAs) that inhibit the reverse transcription of pgRNA to rcDNA. However, these therapies do not directly target the cccDNA reservoir [4] and viremia rebounds once treatment is stopped, even when peripheral levels of viral DNA have remained undetectable for months or years [5]. There is a growing impetus to identify curative therapies for HBV [6]. Despite its central role in the HBV life cycle, our understanding of the viral and host factors that regulate cccDNA abundance and half-life is limited[7]. cccDNA half-life can be defined as the time for the number of copies in the liver to reduce by half and will depend on a number of factors, including cccDNA ‘lifespan’ (the time an individual cccDNA molecule persists) and the genesis of new cccDNA via extra-cellular virus infection or intra-cellular amplification [8,9] (Fig.1a). A recent *in vitro* study reported a 40 day half-life of HBV cccDNA in infected HepG2-NTCP cells [2], with an estimated lifespan of 58 days. However, the cccDNA lifespan in the human liver is unknown.

**Fig 1:**
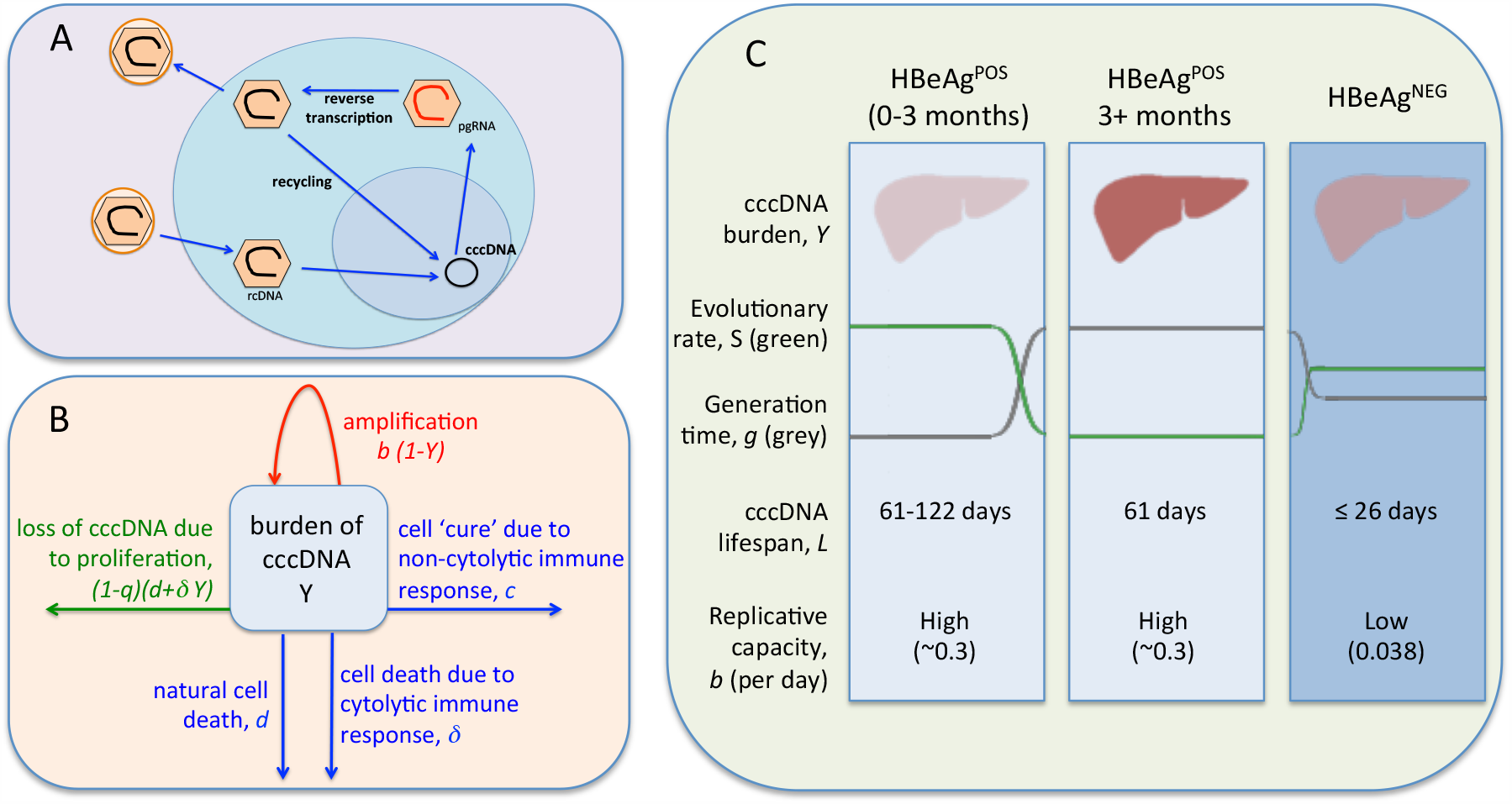
Summary of the HBV life cycle, mathematical model, and estimated cccDNA lifespan. **A:** Simplified HBV replication cycle. A virus particle containing relaxed circular DNA (rcDNA) enters a hepatocyte (blue circle) and is uncoated. The rcDNA is transported to the nucleus (purple circle) and repaired to generate cccDNA. This cccDNA is the transcriptional template for all viral RNAs, including pre-genomic (pgRNA), which is transported to the cytoplasm, encapsidated, and converted into rcDNA by error-prone reverse transcription. The encapsidated rcDNA can be transported back into the nucleus to form more cccDNA (intra-cellular amplification), or enveloped and released as virions that can infect hepatocytes (extra-cellular amplification). **B**: Structure of the mathematical model. This is a single compartment model representing the burden of cccDNA in the liver, Y, over the course of infection. The cccDNA burden can increase due to amplification (intra- and extra-cellular), where *b* is a measure of the within-host replicative capacity of cccDNA. cccDNA can be cleared from the liver due to natural cell death, at rate *δ* per day, cytolytic immune responses at rate *δ* per day, and non-cytolytic immune responses at rate *c* per day. Proliferation can also result in loss of cccDNA at rate *(1-q)(d+δY)*, where *q* is probability that an individual cccDNA survives mitosis. **C**: Representation of the model dynamics and key results, where the numbers give the most likely values inferred by fitting the mutation and evolutionary rates to the model. The darker the colours on the figure the higher the cccDNA burden (reds) and the stronger the immune response (blues).

The lifespan of cccDNA molecules most likely changes over the course of CHB, influenced by host and viral factors [10], including the rate of hepatocyte proliferation [11,12]. The natural history of CHB is often classified by four clinical or virological phases of infection and/or hepatitis (according to European Association for the Study of the Liver 2017[13]). However, it is increasingly recognised that this description fails to capture the diversity of viral replication in the liver and misleadingly suggests a linear progression over time from one phase to the next. To avoid difficulties associated with this rigid phenotypic structure of disease, and to focus on viral dynamics rather than disease pathology, we instead consider two states: HBeAg^POS^ that is associated with high peripheral HBV DNA levels (viral load - VL), and HBeAg^NEG^, which is associated with lower VLs [14]. Early in infection cccDNA is transcriptionally active and translation of pre-core/pgRNA results in detectable levels of hepatitis B e antigen (HBeAg) in the periphery, associated with high VL [15]. In later stages of infection after HBeAg seroconversion there is a loss of HBeAg that associates with more efficient immune targeting of infected cells [16], leading to a reduction in VL and a shortening of cccDNA lifespan. The higher hepatocyte death rates during HBeAg^NEG^ CHB infection will induce hepatocyte proliferation [15]. The extent to which cccDNA is lost during hepatocyte mitosis is uncertain [9], but unless all cccDNA episomes survive mitosis, the increased proliferation rate of infected cells will shorten the average lifespan of cccDNA [12,17,18].

Double-stranded DNA viruses typically have low mutation rates, but since rcDNA is generated via error prone reverse transcription in the *hepadnaviridae*, they have higher mutation rates than other DNA viruses (Fig 1A) [19]. The estimated mutation rate for avian hepadnavirus is 2×10^−5^ substitutions per site per infected cell (s/s/c) [20], and an upper limit of 8.7×10^−5^ s/s/c has been estimated for HBV [21], similar to estimates for RNA retroviruses [22]. The evolutionary rate measures how quickly mutations become fixed in a population over a period of time [23]. Strikingly, the evolutionary rate of HBV is much lower than for RNA viruses with similar mutation rates [24–26]. Different mechanisms could explain this observation, including the biological constraint of multiple overlapping reading frames in the HBV genome [24], limited viral population size in the liver, or long cccDNA lifespan [24,25]. Both evolutionary constraint and population size should only influence the rate of evolution of variants that experience selection [27]. For neutral or near neutral mutations, the long cccDNA lifespan provides the simplest explanation for the low evolutionary rate of HBV [25].

We propose the within-host evolutionary rate of HBV can be used to estimate cccDNA lifespan and the replicative capacity (a combined measure of extra-cellular viral production and intra-cellular amplification) of the virus. We developed a novel mathematical model that uses published HBV mutation and evolutionary rates [20,24,26], to infer the lifespan and replicative capacity of cccDNA during different phases of CHB. Although we use our model to consider the neutral rate of evolution, it is essentially an ecological model, in that we do not assume any viral phenotypic evolution. To the best of our knowledge, these are the first estimates of cccDNA lifespan in treatment naïve subjects and provide important insights into the HBV reservoir that will be valuable for the design and evaluation of future treatment interventions.

## RESULTS

We developed a mathematical model describing the number of cccDNA molecules in the liver that is independent of infected cell frequency, and accounts for intra- and extra-cellular cccDNA amplification and loss of cccDNA during hepatocyte mitosis (Fig 1B and Methods, Eqs 1 and 2). Using this model we derived expressions for the viral generation time, defined as the typical time for one cccDNA molecule to generate another cccDNA molecule at time *t* since infection, *g(t)* (Eq 5), and the neutral rate of evolution at time *S(t)* (Eq 6). At equilibrium, we show that the lifespan of cccDNA, *L*_*E*_, is equal to the virus generation time, *g*_*E*_, which is given by the neutral mutation rate divided by the neutral rate of evolution, *μ /S*_*E*_ (Eq 8). The notation used throughout is given in Table 1.

**Table 1:** Notation used to describe the model.

|  |  |
| --- | --- |
| $N(t), N_E$ | Number of cccDNA molecules at time $t$ since infection and at equilibrium |
| $K$ | Maximum possible number of cccDNA molecules |
| $Y(t), Y_E$ | cccDNA burden in the liver at time $t$ since infection and at equilibrium, expressed as a proportion of the maximum possible number of cccDNA, $Y(t)=N(t)/K$, $Y_E= N_E/K$ |
| $\rho(t)$ | Per capita proliferation rate of cccDNA at time $t$ since infection |
| $g(t), g_E$<br>$g_{Y \ll 1}$ | Generation time of cccDNA at time $t$ since infection, at equilibrium, and when few cells are infected |
| $S(t), S_E$ | Neutral rate of evolution at time $t$ since infection and at equilibrium |
| $L_E, L_{Y<<1}$ | Life span of cccDNA molecules at equilibrium and when few cells are infected |
| $\beta$ | Per capita replicative capacity, defined as the per capita growth rate of cccDNA when few cells are infected and in the absence of infected cell death or loss of cccDNA due to non-cytolytic immune responses. |
| $R_0$ | The basic reproductive number of cccDNA (the number of cccDNA molecules a single cccDNA produces in its lifetime in an otherwise uninfected population of hepatocytes) |
| $b = \beta K$ | Replicative capacity of cccDNA (a rescaled measure of the per capita replicative capacity) |
| $d$ | Per capita natural death rate of hepatocytes |
| $\delta$ | Per capita additional death rate of infected hepatocytes due to cytolytic immune responses |
| $c$ | Per capita loss rate of cccDNA due to non-cytolytic immune responses |
| $q$ | Probability that a cccDNA molecule survives mitosis |
| $\mu$ | Mutation rate of cccDNA (substitutions per site per reproduction) |

<sup>+</sup> Throughout we use days as the time unit and all rates are per day.

### Lifespan of cccDNA

From the published estimates for the mutation [20] and evolutionary rates of HBV [24,26], we inferred the probability distributions for cccDNA lifespan during HBeAg^POS^ and HBeAg^NEG^ phases of CHB using Eq 8. The predicted lifespan of cccDNA during HBeAg^POS^ infection, and when VLs are high and stable, is 61 days (5^th^-95^th^ percentiles 36-236 days; Fig 2, orange line). In contrast, during HBeAg^NEG^ infection the lifespan of cccDNA is estimated at only 26 days (16-81 days; Fig 2, blue line).

**Fig 2.**
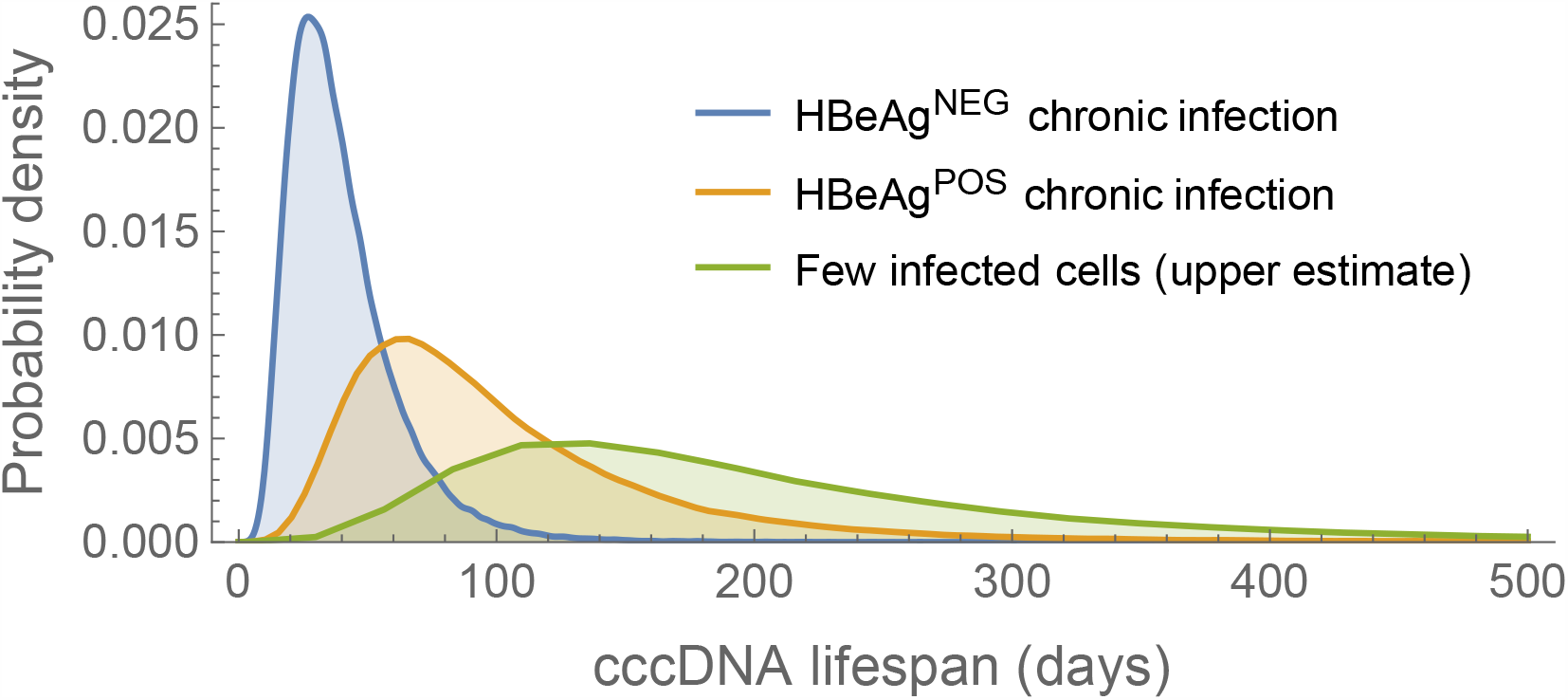
Probability distributions for cccDNA lifespan during different stages of HBV infection. The distributions for cccDNA in stable HBeAg^POS^ and HBeAg^NEG^ chronic infection are based on the neutral mutation rate and rate of neutral evolution (orange and blue lines, respectively). If the cccDNA burden during HBeAg^NEG^ infection is not stable, but gradually falling (i.e. the basic reproduction number, *R*_*0*_, is less than one) the lifespan will be slightly less than inferred here. The upper estimate reflects the maximum likely cccDNA lifespan when few cells are infected, based on the neutral rate of evolution during HBeAg^POS^ infection and assuming no cccDNA survives mitosis (*q*=0; green line).

The shorter lifespan of cccDNA during HBeAg^NEG^ compared to HBeAg^POS^ infection can be explained by higher rates of cccDNA clearance (Eq 9). This may reflect changes in the immune environment due to HBe-antigen seroconversion that is associated with increased cytolytic and non-cytolytic immune responses (*δ* and *c* respectively). Increased host immune responses during HBeAg^NEG^ infection could push the basic reproduction number, *R*_*0*_, of cccDNA below one (Eq 3) due to the higher clearance rates of cccDNA molecules, and also due to reduced replicative capacity, *b*, of cccDNA. If *R*_*0*_<1, the number of cccDNA will not reach a stable level but will continually decline. In this non-equilibrium situation the lifespan of cccDNA may be less than our inferred 26 days since the viral generation time will be greater than the lifespan of cccDNA (Fig 3 and methods).

**Fig 3.**
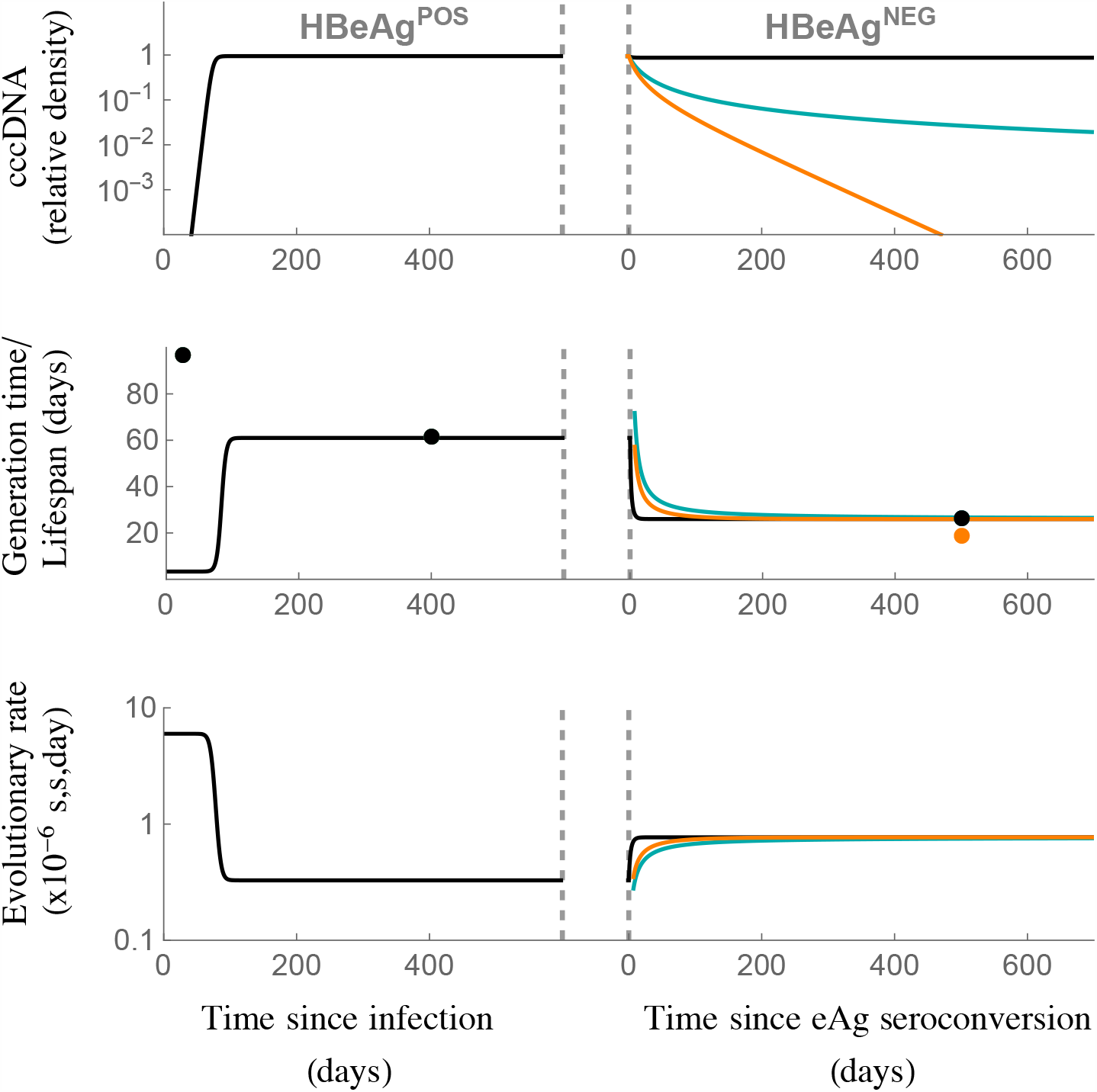
Population and evolutionary dynamics of cccDNA for the within-host model. We assumed no cccDNA survives mitosis (*q=0;* see S1 Fig for the case where *q*=1). The model was parameterised assuming the generation time, *g*, during chronic HBeAg^POS^ infection is 61 days, and during chronic HBeAg^NEG^ infection is 26 days, in line with our predictions for cccDNA generation time *in vivo*. The top panel shows cccDNA burden, where 1 represents the maximum. The middle panel shows the viral generation time (lines) and cccDNA lifespan during key stages of infection (dots, derived from Eqs 9 and 10). The bottom row shows the evolutionary rates. Black line: replicative capacity during HBeAg^NEG^ infection remains the same as during HBeAg-postive infection (*b*_*eAg+*_*= b*_*eAg-*_*=0*.*3* per day). Blue line: replicative capacity falls to *b*_*eAg-*_=0.038 per day during HBeAg^NEG^ infection, and *R*_*0*_=1. Orange line: replicative capacity falls to *b*_*eAg-*_*=0*.*038* per day and *R*_*0*_=0.7. See Table 1 for all other parameters.

Our model suggests cccDNA lifespan can be up to two times longer when few cells are infected compared to when most cells are infected cells (see methods; compare Eqs 9 and 10). When few cells are infected there is less cell death due to cytolytic immune responses, a lower rate of hepatocyte proliferation to maintain the number of hepatocytes, and consequently reduced loss of cccDNA via mitosis of infected cells. This is of more than theoretical interest, because when estimating how long it will take to deplete the cccDNA reservoir on treatment, it is the lifespan of cccDNA when relatively few cells are infected that is important since treatment is known to reduce the cccDNA load. The maximum expected cccDNA lifespan, corresponding to HBeAg^POS^ infection, few infected cells, and no cccDNA surviving mitosis, is 123 days (71-472 days; Fig 2, green line). Reports for duck hepatitis B virus (DHBV) show a high proportion of cccDNA survives mitosis [18]. In contrast, for HBV recent experimental [9,17] and modelling [12] results suggest that relatively few cccDNA molecules survive mitosis, making this longer lifespan a reasonable expectation.

### Dynamics of the mathematical model

To demonstrate the behaviour of our model we present examples of the dynamics when no cccDNA survives mitosis (q=0, Fig 3; see S1 Fig for model dynamics when *q*=1). We used parameters that are compatible with our estimated cccDNA generation times (61 days during HBeAg^POS^ infection and 26 days during HBeAg^NEG^ infection). Since hepatocytes are long-lived we defined the natural death rate as *d*=0.002 per hepatocyte per day throughout and, for simplicity, we set *c*=0 under the assumption that cytolytic responses have greater antiviral activity than non-cytolytic responses. We assume a neutral mutation rate *μ* =2×10^−5^ s/s/c [20]. The model dynamics when *q=1* are similar to the case where *q=0*, apart from the lifespan of cccDNA in the early stages of infection is predicted to be higher if q=1 (see below). A graphical representation of the results is given in Fig 1C, and a summary of the parameters in Table 2.

**Table 2:** Parameters used to describe the model dynamics^1,2^.

|  | Model 1 |  | Model 2 |  |
| --- | --- | --- | --- | --- |
|  | *No cccDNA survives mitosis* |  | *All cccDNA survives mitosis* |  |
|  | HBeAg<sup>POS</sup> | HBeAg<sup>NEG</sup> | HBeAg<sup>POS</sup> | HBeAg<sup>NEG</sup> |
| $q$ | 0 | 0 | 1 | 1 |
| $g$ | $g_E=61$ | $g_E=26$<br>${}^3g_{Y<<1}=26$<br>${}^3g_{Y<<1}=26$ | $g_E=61$ | $g_E=26$<br>${}^3g_{Y<<1}=26$<br>${}^3g_{Y<<1}=26$ |
| $R_0$ | 30.0 | 13.6<br>1.0<br>0.7 | 37.5 | 7.9<br>1.0<br>0.7 |
| $b$ | 0.3 | 0.3<br>0.038<br>0.038 | 0.3 | 0.3<br>0.038<br>0.038 |
| $d$ | 0.002 | 0.002 | 0.002 | 0.002 |
| $\delta$ | 0.006 | 0.018<br>0.034<br>0.050 | 0.014 | 0.036<br>0.036<br>0.052 |
| $c$ | 0 | 0 | 0 | 0 |
| $\mu$ | $2 \times 10^{-5}$ | $2 \times 10^{-5}$ | $2 \times 10^{-5}$ | $2 \times 10^{-5}$ |

<sup>1</sup>All rates are given per day, and generation times are listed as days
<sup>2</sup>Where three values are given these refer to the alternative parameters used for the different trajectories presented in Fig 3 and S1 Fig (top parameter, black line; middle parameter, blue line; bottom parameter, orange line).
<sup>3</sup>Since $R_0 \leq 1$ , a non-zero equilibrium is not reached.

#### HBeAg^POS^ infection

The replicative capacity of cccDNA, *b*, was chosen to be 0.3 per day so that the peak number of cccDNA molecules in the liver is reached at approximately 3 months since infection, in line with reported observations [28]. The per capita death rate of infected cells due to cytolytic immune responses, *δ*, was determined assuming a cccDNA generation time at equilibrium of 61 days, and solving Eq 9 for *δ* (giving *δ*=0.006 per day if *q=0;* the associated *R*_*0*_ is 30).

Under these assumptions, during the first few months of infection the cccDNA burden (number of cccDNA divided by the maximum number of cccDNA) increases rapidly, leading to a short viral generation time predicted by the model of 3.3 days (Eq 11, Fig 3, S1 Fig). A recent study estimated an eclipse period of approximately 3 days for a newly infected cell to produce viral particles [2], so our estimated viral generation time seems reasonable. This short generation time of cccDNA during early infection contrasts with the long cccDNA lifespan (dots in Fig 3, S1 Fig; note the longer lifespan predicted if *q=0* compared to *q*=1). The neutral rate of evolution is also predicted to be high during this early stage of infection due to the short generation time.

As infection progresses, the viral generation time increases due to fewer susceptible target cells (Eq 5), in line with results in epidemiology [29], and this in turn reduces the evolutionary rate (Eq 6). This dependency of evolutionary rate on epidemiological dynamics has been noted in a previous simulation study on within-host viral infection [30], but is generally an underappreciated factor influencing evolutionary rates. At equilibrium, the estimated viral generation time and cccDNA lifespan are the same, and it is this equivalency that enables us to determine these parameters from the neutral rate of evolution, independent of the parameters of the model (see Methods). Due to the long lifespan of infected hepatocytes, a high cccDNA burden is reached in the model. This is in line with observations that most hepatocytes are infected at peak infection [31].

#### HBeAg^NEG^ infection

We assumed the transition from HBeAg^POS^ to HBeAg^NEG^ occurs after an arbitrary amount of time after HBeAg^POS^ equilibrium is reached and associates with a reduced cccDNA generation time from 61 to 26 days. If this reduced generation time is not accompanied by a decrease in replicative capacity, only a modest fall in the cccDNA burden is predicted (Fig 3, blue line; 5=0.018 per day and *b*=0.3 per day). However, this is inconsistent with *in vivo* infections, where the number of cccDNA molecules and extracellular HBV DNA levels (VL) typically decline by several orders of magnitude after transition to HBeAg^NEG^ infection [32,33]. If the cccDNA burden is low (Y<<1), then replicative capacity, *b*, is estimated by the reciprocal of the generation time (*b=1/g;* Eq 5). For a generation time of 26 days, this gives *b*=0.038 per day, leading to an estimated 10-fold reduction in the ability of cccDNA to reproduce during HBeAg^NEG^ compared to HBeAg^POS^ infection. In Fig 3, the orange line shows the model dynamics given this decline in *b*, and when *R*_*0*_*=1* during HBeAg^NEG^ infection (i.e. *δ* = 0.034 per day and *b*=0.038 per day). In this case, the cccDNA burden falls at a relatively modest rate. Perhaps more likely is that *R*_*0*_ < 1 and the number of cccDNA molecules continues to decline. The green line shows the dynamics if *R*_*0*_=0.7 (*δ* = 0.050 per day). However, even with this modest increase in *δ*, the number of cccDNA is predicted to fall rapidly.

The difficulty in explaining low but steady VL using standard within-host virus models, and the sensitivity of VL to model parameters when *R*_*0*_ is close to one, have been acknowledged previously, particularly in relation to HIV-1 infections [34–36]. Possible explanations for the low numbers of cccDNA during HBeAg^NEG^ infection and low rates of spontaneous cure include the existence of a small number of hepatocytes that are susceptible to infection, resulting in low numbers of cccDNA molecules even if *R*_*0*_ is high [34], or the existence of a metapopulation-type partitioned structure in the liver, which enables the cccDNA to persist when *R*_*0*_ is low [36].

### Estimated time to eradicate cccDNA on treatment

When few cells are infected, the inferred cccDNA lifespan is 123 days during HBeAg^POS^ infection if *q=0*. Even with this longer estimate for cccDNA lifespan, if there are 10^12^ cccDNA molecules at the start of treatment (see methods), we would expect the reservoir to be depleted after less than ten years of treatment (Eq 13, Fig 4A). Moreover, if treatment is initiated during HBeAg^NEG^ CHB the time to eradicate cccDNA is predicted to be even faster (only 1.5 years) with a lifespan of 26 days, and a lower number of cccDNA molecules (2×10^9^) in the liver at the start of treatment. However, these predictions are in stark contrast to what is observed in the clinic, where a high proportion of individuals remain infected after many years of continuous treatment [37] and there is no appreciable difference in treatment mediated cure in HBeAg^NEG^ or HBeAg^POS^ patients [8,38]. The discrepancy may arise due to our estimated cccDNA lifespan being too short. An estimated lifespan of 236 days during HBeAg-postive CHB still lies within our 95% confidence interval, and would give a time to eradication, and hence sterilizing cure between 18 and 36 years (Eq 13). However, this does not explain the long time to eradicate cccDNA during HBeAg^NEG^ infection. Alternative explanations include ongoing (albeit reduced) cccDNA amplification during NA treatment (*b*>0) [8,39], or the presence of a long-lived subset of infected hepatocytes [17,40].

**Fig 4.**
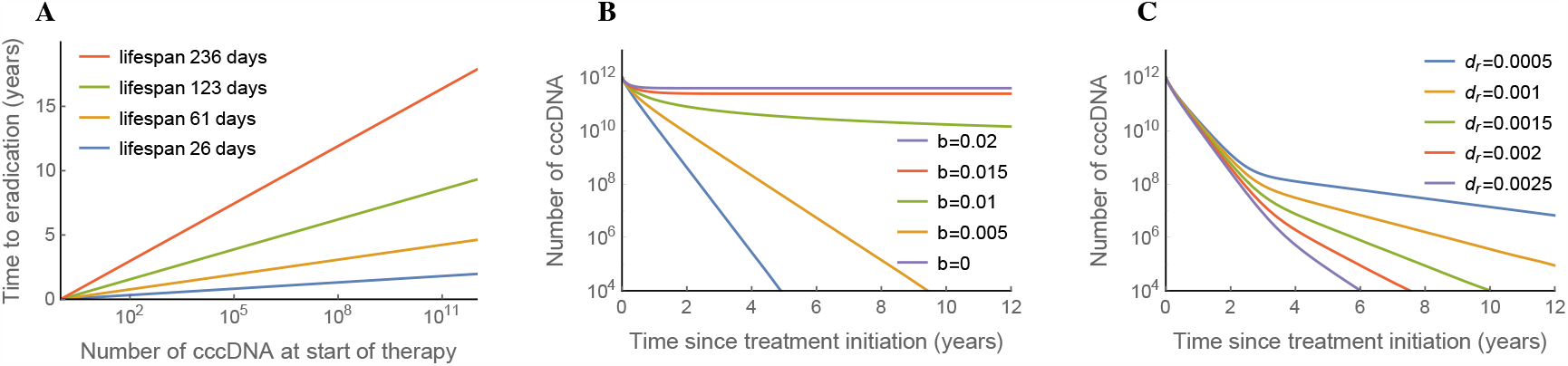
Effect of NA treatment predicted by the model. A: Predicted time for cccDNA to be eradicated in the absence of any cccDNA reproduction (*b*=0). B: cccDNA dynamics whilst on treatment, assuming some residual reproduction. For all runs *d*=0.002 per day, *δ*=0.006 per day, *c*=0, *q=0*. C: cccDNA dynamics on treatment, assuming no residual reproduction (*b*=0) but 0.1% cccDNA is long-lived, for different death rates of long-lived cells, *d*_*r*_ per day. For normal cells *d*=0.002 per day, *δ*=0.006 per day,*c*=0, *q*=0, and for long-lived cells *δr*=0, *c*_*r*_=0, *q*=0. The maximum number of cccDNA was assumed to be 10^12^, and all model runs were started at equilibrium in the absence of treatment (*b*=0.3 per day).

To evaluate these two scenarios, we modelled cccDNA dynamics in CHB patients on treatment assuming different levels of viral replication (Fig 4B) or a subset of long-lived cells (Fig 4C, S2 Fig). The dynamics of cccDNA are sensitive to the amount of replication, making it unlikely that ongoing amplification alone explains the failure of treatments to eliminate cccDNA. Apart from a narrow range of replicative capacities, either a high and steady cccDNA burden, or relatively rapid cccDNA elimination, is predicted on treatment. The existence of a long-lived population of infected hepatocytes is more robust to differences in model parameters, with a gradual increase in the time to eradicate cccDNA as the death rate of long-lived cells is increased, making a long-lived population a more parsimonious explanation for the slow decline in the HBV reservoir. However, since the decay dynamics of the reservoir on treatment can be complex, and differ between individuals [39], a combination of factors most likely explains the clinical observations.

## DISCUSSION

We provide a new model to estimate the HBV cccDNA lifespan based on reported mutation and within-host evolutionary rates [20,24,26]. The lifespan of cccDNA is an important component of the half-life of the cccDNA reservoir, which describes how the population of cccDNA molecules in an individual declines over time. We predict an average cccDNA lifespan of 61 days during HBeAg^POS^ CHB compared to only 26 days in the HBeAg^NEG^ phase of infection. Although estimates for the mutation and evolutionary rates for HBV are associated with high levels of uncertainty, our predicted lifespan is in agreement with *in vitro* studies showing a 40 day half-life of HBV cccDNA [2] and an estimated half-life of 33-57 days in woodchucks and ducks *in vivo* [41,42]. As far as we are aware, this is the first time cccDNA lifespan has been estimated during untreated infection. The lower lifespan during HBeAg^NEG^ infection is consistent with a study in which VL data during therapy was fitted to a mathematical model, concluding that the turnover of infected cells is higher if therapy is initiated during HBeAg^NEG^ infection [16], although our predictions for cccDNA persistence are longer [16].

The shorter cccDNA lifespan during HBeAg^NEG^ CHB may reflect host immune responses, with our model suggesting a doubling of the clearance rate compared to HBeAg^POS^ infection. However, this increased clearance rate is predicted to have a modest effect on the total number of cccDNA molecules. As well as inferring the lifespan of cccDNA, we inferred cccDNA replicative capacity (a combined measure of intra and extracellular amplification). Our results predict an approximate ten-fold reduction in replicative capacity between HBeAg^POS^ and HBeAg^NEG^ phases of infection, which may reflect host innate or adaptive immune responses suppressing viral production and/or intra-cellular amplification. This can explain the lower cccDNA levels reported in HBeAg^NEG^ CHB [32,33], and is consistent with observations that the replicative capacity of cccDNA in the HBeAg^NEG^ phase of infection is reduced compared to HBeAg^POS^ infection [32]. This may reflect immune control at the level of the viral epigenome, but without cell death [43].

In our modelling approach, we have assumed that viral dynamics are driven by host-factors, in the absence of viral phenotypic evolution. The HBeAg^NEG^ phase of infection is often associated with an emergence of precore mutations that may affect HBV replication and stability of cccDNA [44,45]. Viral phenotypic evolution is likely to have an important role in viral dynamics, and we speculate that these viral phenotypic changes could increase the rate of intra-cellular amplification, but at the expense of killing the host cell, and therefore could represent a form of intra-host short-sighted evolution [25].

Our estimates for cccDNA lifespan have implications for curative treatment strategies. If NA therapy inhibits all cccDNA amplification, we would predict HBV to be cured after 1 to 10 years of continuous treatment. However, this is not observed in the clinic, with only 1% of individuals clearing HBsAg each year [37]. Possible explanations for this discrepancy are that NAs do not inhibit all intra- and extra-cellular amplification [8,39], or the existence of long-lived infected cells [17,40]. Our model is consistent with the presence of long-lived infected cells providing the most parsimonious explanation for sustained infection on treatment. There is growing evidence that there is negligible intra-cellular cccDNA amplification in human HBV infection [9], and since NA treatment will inhibit the genesis of viral particles this will prevent extra-cellular amplification. Furthermore, the dynamics of cccDNA clearance is sensitive to the assumed amplification rates, and therefore if amplification alone explains the dynamics we would expect to see a proportion of individuals clearing infection within 1-2 years of starting treatment. The presence of long-lived HBV infected cells has parallels with the HIV reservoir, where long-lived latent-infected CD4^+^ T cells prevent cure [46]. Distinguishing between residual amplification and long-lived infected cells will help define the expected impact of treatment strategies that prevent cccDNA replication, compared to those directly targetting cccDNA. As HBV evolution will only occur if there is cccDNA amplification, it may be possible to distinguish between these two mechanisms by measuring the rate of cccDNA evolution whilst on treatment.

Our estimates of cccDNA persistence and amplification provide insights into mechanisms underlying CHB and will inform our understanding of how spontaneous or therapeutic clearance may be achieved. Given different infection profiles among individuals, and limited datasets available for our model, the confidence intervals of our estimations are wide. Our analysis exemplifies the power of modelling as a tool to inform therapeutic interventions and highlights the need for genomic studies to determine HBV evolutionary rates in CHB.

## METHODS

To derive estimates of HBV cccDNA lifespan using the neutral mutation rate and the rate of evolution we developed a deterministic mathematical model describing the dynamics of cccDNA during the course of treatment naïve CHB. We used this model to derive expressions for viral generation time and neutral rate of evolution, both of which are predicted to change during the course of infection. Finally, we derived expressions for the lifespan of cccDNA during (i) stable CHB and (ii) when the proportion of infected cells is low, as would be expected in early stages of infection or in the first few months of NA treatment.

### A within-host model of HBV dynamics

HBV cccDNA can replicate via intra-cellular and extra-cellular routes (Fig 1A), with a reported copy number between 1-50 molecules within a single hepatocyte nucleus [2,17,31,47–49] (the higher estimates tend to be for DHBV and lower estimates for human HBV [9]). Since cccDNA can be lost during mitosis, we modelled the number of cccDNA copies in the liver, rather than the number of infected cells. To do this, we implicitly assume that viral production is proportional to the number of cccDNA molecules. This is a reasonable assumption since VL has been reported to associate with increasing cccDNA copy numbers [32,50].

We describe the number of copies of cccDNA in the liver at time *t* since infection, *N(t)* as:

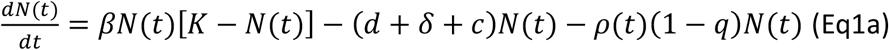

where the first term describes the increase in cccDNA due to intra- and extra-cellular amplification. We assume that the rate of increase is density dependent, with a maximum per capita growth rate *β* per day and a maximum possible number of cccDNA, *K*. We assume *K* is constant since proliferation ensures the number of hepatocytes in the liver remains stable during infection [15], and because evidence suggests there is a maximum number of copies of cccDNA that can persist within each hepatocyte, which is virally controlled [50-52]. In reality, the value of K is likely to fall during infection, for example due to sections of the liver becoming cirrhotic and/or if hepatocytes expressing HBsAg from integrated viral DNA have reduced susceptibility of infection. However, since ours is a density dependent model, this will have only a small effect on model dynamics if the change in K is much slower than viral dynamics, as is expected. Moreover, our estimates for cccDNA lifespan (below) are based on equilibrium values and are therefore independent of the value of K.

The second term describes the rate at which cccDNA is lost due to the natural death of hepatocytes and the host immune response, under the assumption that cccDNA is randomly distributed among infected hepatocytes. We assume that hepatocytes, and therefore cccDNA, have a natural per capita death rate *d* per day. Infected hepatocytes (and hence cccDNA) have an additional per capita death rate *δ* per day due to cytolytic immune responses, and cccDNA is lost at per capita rate *c* per day due to non-cytolytic immune responses.

The final term describes the loss of cccDNA due to cell proliferation. Uninfected and infected hepatocytes are assumed to proliferate at the per capita rate *ρ*(*t*) per day, and hence cccDNA will be exposed to proliferation at rate, *ρ*(*t*), with a probability *q* that a cccDNA molecule will survive mitosis. Since the maximum possible number of cccDNA, *K*, is assumed to be constant, proliferation and cell death are balanced, hence:

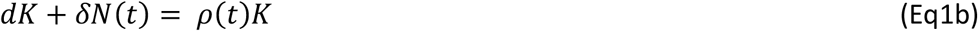

Here, the left-hand side represents the loss in carrying capacity due to the natural death rate of all hepatocytes (including infected hepatocypes), and the additional death rate incurred by infected hepatocytes as a direct result of infection. We are making the assumption that cccDNA are randomly distributed among infected hepatocytes, and that the natural death rate of hepatocytes is independent of whether they are infected or not. Thus, for example, a 5% drop in the number hepatoctyes results in a 5% drop in cccDNA carrying capacity. The right-hand side of the equation represents consequential gain in cccDNA carrying capacity due to proliferation that is needed for K to remain constant.

A complete expression for the dynamics of *N(t)* can be found by solving Eq 1b for *ρ* (*t*) and substituting into Eq 1a. To simplify further, we consider the cccDNA burden in the liver, *Y*(*t*) *= N*(*t*)*/K*, rather than the total number of cccDNA molecules, giving us:

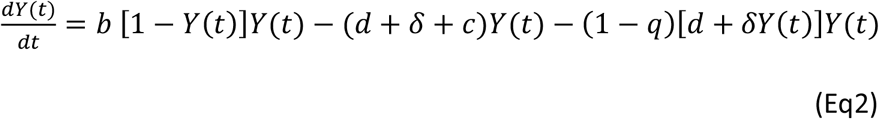

where *b* = *βK* is a rescaled measure of cccDNA replicative capacity. From this equation we can calculate the basic reproductive rate of cccDNA, *R*_*0*_, which is defined as the number of new cccDNA molecules a single cccDNA molecule will produce in a susceptible population of hepatocytes:

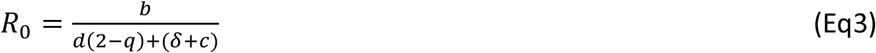

If R_0_<1, then the infection cannot be sustained in the long term. At equilibrium, the cccDNA burden is given by:

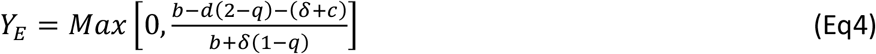

which is equivalent to the cccDNA burden during stable chronic infection. Our model considers the number of cccDNA molecules independent of their distribution within cells. This is similar to the “single copy” modelling assumption used in [18], in which only one cccDNA molecule can persist in a cell, and which was shown to produce almost identical dynamics to one in which multiple copies of cccDNA are explicitly modelled within infected cells [18].

### An expression for the neutral rate of HBV evolution

In a large well-mixed viral population, and in the absence of selection, the rate of evolution at time *t* is given by *S*(*t*) = *μ/g* (*t*), where *μ*. is the (neutral) mutation rate, measured per site per viral generation, and *g(t)* is the generation time [53]. For our within-host model of HBV infection, *g* is equivalent to the typical amount of time it takes for one cccDNA molecule to replicate another molecule. This is similar to the meaning of the generation time in demography and epidemiology [29,54,55]. From Eq 2, we can calculate the ‘backwards’ generation time as:

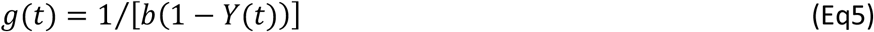

At time *t* since initial infection, the neutral substitution rate is therefore given by:

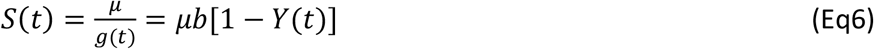

Since intra- and extra-cellular amplification involve an error-prone reverse transcription step (in both cases cccDNA is the template for producing pgRNA, which is then reversed transcribed to form rcDNA), we have assumed they have similar mutation rates. Substituting *Y*_*E*_ into Eq 6, we can find an expression for the neutral rate of evolution rate at equilibrium:

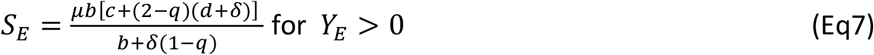

### Lifespan of cccDNA during steady state infection

In our model, at equilibrium the generation time of HBV will be equal to the typical cccDNA lifespan, *L*_*E*_. At equilibrium the number of cccDNA molecules remains constant, and therefore the rate at which cccDNA is produced is equal to the rate at which cccDNA is lost due to infected cell death, non-cytolytic clearance of cccDNA, and proliferation of infected cells. Since the reciprocal of the production rate is equal to the generation time, and the reciprocal of the rate cccDNA is lost is the typical lifespan of cccDNA, at equilibrium, viral generation time and cccDNA lifespan are identical (*g*_*E*_ *= L*_*e*_). This relationship holds because of our assumption of constant death rate and hence exponentially distributed lifetimes of cccDNA [29]; see [29,54] for how this changes for different distributions.

Using the equivalence of *g*_*E*_ and *L*_*E*_, the lifespan of cccDNA at equilibrium can be determined from the mutation and neutral evolution rates by rearranging the first part of Eq 6:

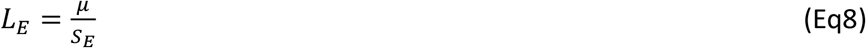

Substituting the expression for *S*_*E*_ from Eq 7 into Eq 8, we can write an expression for the lifespan of cccDNA at equilibrium based on the model parameters:

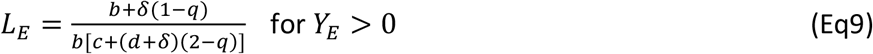

### The lifespan of cccDNA when few cells are infected

If infection increases the death rate of hepatocytes, then the level of proliferation (to replace eliminated cells) will be larger the more cells are infected. Consequently, the lifespan of cccDNA when few cells are infected (e.g. during early phases of infection or during spontaneous clearance of infection, or after prolonged successful suppressive treatment) may differ from the lifespan during HBeAg^POS^ or HBeAg^NEG^ steady state infection. By setting *Y<<1* in equation 2, we can derive an expression for cccDNA lifespan when the copy number or burden is low:

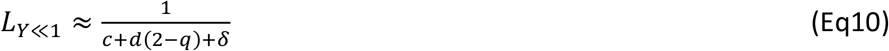

Comparing the expressions for *L*_*E*_ and *L*_*Y<<*1_, we can see that if all cccDNA survives mitosis (*q*=*1)* or infection has a minimal effect on the death rate of infected cells (*δ* =0), then cccDNA lifespan remains unchanged during infection (as long as *d* and *c* don’t change). However, if these conditions are not met, then the lifespan of cccDNA when few cells are infected, *L*_*Y<<*1_, can be up to double the lifespan during chronic stable infection, *L*_*E*_, for identical model parameters (e.g. when *q=c=d=0*, and *b>> δ)*.

As we noted above, the cccDNA lifespan is only equivalent to the generation time at equilibrium. Using equation 5, when few cells are infected, the generation time is given by:

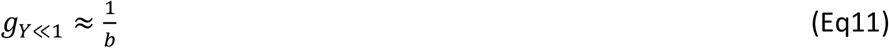

This has also been observed in the epidemiological literature [29]. Combining equations 3, 10 and 11 we see that:

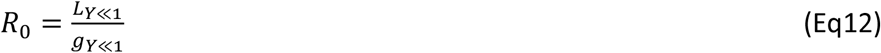

If *R*_*0*_ >1 and few cells are infected (i.e. the number of cccDNA is increasing) the life expectancy of cccDNA will be greater than the viral generation time, whereas if *R*_*0*_ *< 1* the life expectancy will be less than the viral generation time. This might be the case if, for example, increased immune responses associated with HBeAg^NEG^ infection push *R*_*0*_ below one.

### Estimating the generation time and lifespan of cccDNA from within-host evolutionary rates

During stable chronic infection, the lifespan of cccDNA, *L*_*E*_, equals the viral generation time, *g*_*E*_, with *g*_*E*_ = *μ/S*_*E*_ (Eq 6). Although the mutation rate of HBV has not been determined, for avian hepadnavirus it has been estimated at 2×10^−5^ s/s/c (in the range 0.8×10^−5^ to 4.5×10^−5^; [20]). Since we are interested in the neutral rate of evolution, we assume that a third of all mutations in non-overlapping reading frames are synonymous, and that synonymous mutations are neutral or nearly neutral [56], giving a neutral mutation rate of around 0.67×10^−5^ s/s/c (0.3×10^−5^ to 1.5×10^−5^) in nonoverlapping reading frames. To incorporate the uncertainty associated with this estimate, we assumed the probability of the true mutation rate is log-normally distributed with mean 10^−5.2^ and standard deviation 10^0.2^.

Using longitudinal HBV sequence data, rates of evolution for non-overlapping regions of the genome were generated using a relaxed clock method, inferring 16.1×10^−8^ (8.1 x10^−8^, 25.5 x10^−8^) substitutions per site per day (s/s/day) for HBeAg^POS^ and 38.9 x10^−8^ (27.2 x10^−8^, 51.5 x10^8^) for HBeAg^NEG^ chronic infection (the numbers in brackets give the 5% and 95% highest posterior density (HPD) intervals; see Table 5 in [24]). In a separate study, using data from [57], the synonymous rate of evolution in nonoverlapping genomic regions was estimated as half of the overall rate of evolution [26]. Assuming synonymous mutations are neutral, and that the ratio of synonymous to nonsynonymous evolutionary rates is constant during infection, we therefore take the neutral within-host rates of evolution during the HBeAg^POS^ and HBeAg^NEG^ phases of infection to be half the rates of evolution reported in [24] for non-overlapping reading frames. This gives a neutral rate of evolution of 8.0×10^−8^ (4.0×10^−8^, 12.7×10^−8^) s/s/day during the HBeAg^POS^ phase, and 19.5 x10^−8^ (13.6 x10^−8^, 25.8 x10^−8^) s/n/day during the HBeAg^NEG^ phase. We assumed the probability distributions of these rates are normally distributed, with the standard deviation calculated using the difference between the estimated rate and the 5% HPD.

We randomly sampled from each of the probability distribution functions (PDFs) for the mutation rate and substitution rates and used these values to calculate the generation time of cccDNA during HBeAg^POS^ and HBeAg^NEG^ CHB. This was repeated 100,000 times, from which the probability distributions for cccDNA generation time during HBeAg^POS^ and HBeAg^NEG^ chronic infection were estimated using the built in SmoothKernelDistribution function in *Mathematica* [58], Assuming the number of cccDNA rapidly reaches equilibrium during HBeAg^POS^ and HBeAg^NEG^ infection, the virus generation time will provide an approximation of the cccDNA lifespan during stable chronic infection (Fig 3).

### Time to cccDNA eradication on treatment

Apart from when treatment is first initiated, the number of infected cells on treatment will be relatively low. Assuming eradication in our model is achieved when fewer than one cccDNA molecule persists, and there is no cccDNA replication whilst on treatment, the time to eradication can be approximated by:

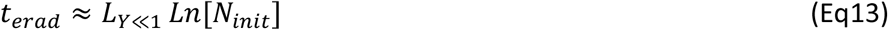

where *N*_*init*_ is the number of cccDNA when therapy is initiated and *Ln* is the natural logarithm. To determine reasonable values for *N*_*init*_, we multiplied the number of hepatocytes in a human liver by the number of cccDNA per hepatocyte during untreated infection. There are about 1.4×10^8^ hepatocytes per gram of human liver [59], and an adult human liver is around 1.5kg, giving approximately 2×10^11^ hepatocytes in total. In a recent study, an average of 6.3 copies of cccDNA per hepatocyte were found during chronic HBeAg^POS^ infection, and 0.01 per hepatocyte during HBeAg^NEG^ infection [32], which gives a total of approximately 1×10^12^ copies of cccDNA during HBeAg^POS^ infection and 2×10^9^ copies of cccDNA during HBeAg^NEG^ infection.

### Model assuming a subset of long-lived hepatocytes

To model cccDNA dynamics in the presence of long-lived hepatocytes, we assume that a constant proportion, *α*, of hepatocytes are long-lived, and the rest ‘standard-lived’, with death of each cell type compensated for by proliferation of that same type. For simplicity, we also assume there is no intra-cellular amplification, and with infection occurring at the same per capita rate in standard- and long-lived uninfected cells. Assuming the cccDNA carrying capacity of standard-lived cells is (1 – *α*)*K*, and of long-lived cells is *αK*, the numbers of cccDNA in standard-lived hepatocytes, *N*, and in long-lived hepatocytes, *L*, are given by:

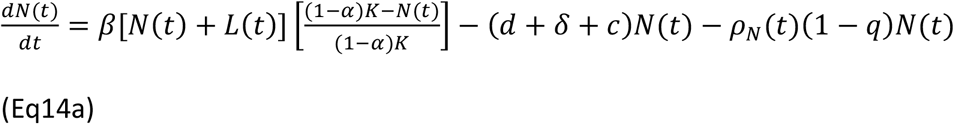

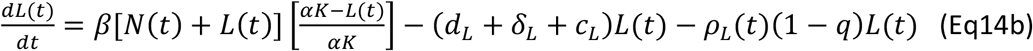

where *d*_*L*_, *δ*_*L*_ and *c*_*L*_ represent the per capita natural death rates, cytoloytic death rate and clearance rates of cccDNA in long-lived cells, and *ρ*_*N*_ and *ρ*_*L*_ the per capita proliferation rates of standard- and long-lived cells. Since deaths of standard (or long-lived) cells are balanced by proliferation of standard (or long-lived) cells, we have the relationships:

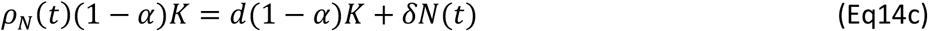

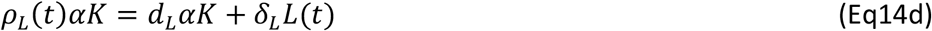

As before, we can substitute expressions for *ρ*_*N*_ (t) and *ρ*_*L*_ (t) found from solving Eqs 14c,d, into Eqs 14a,b, convert numbers into proportions, and simplify, to give expressions for the burden of cccDNA in standard-lived cells, *Y(t)*, and in long-lived cells, *Z(t):*

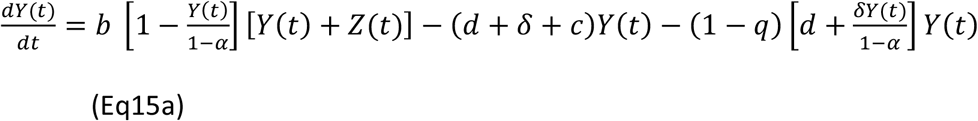

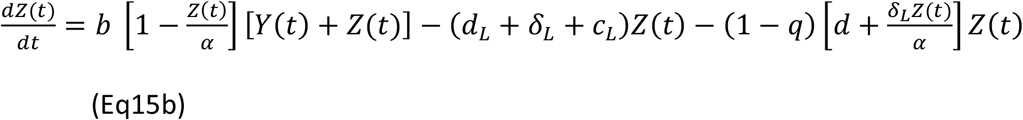

## Data Availability

This paper contains no primary data.

## ACKNOWLEDGEMENTS

Thank you to Fabian Zoulim and Helen Fryer for their helpful comments on earlier versions of this work. KAL and LP are supported by The Wellcome Trust and the Royal Society grant numbers, 107652/Z/15/Z and 202562/Z/16/Z

JAM is supported by EU 2020 Research and Innovation Programme Consortia HEP-CAR under grant agreement No.667273, MRC MR/R022011/1 and The Wellcome Trust IA 200838/Z/16/Z. PM is supported by The Wellcome Trust, grant number 110110/Z/15/Z.

## SUPPORTING FIGURES

**S1 Fig.**
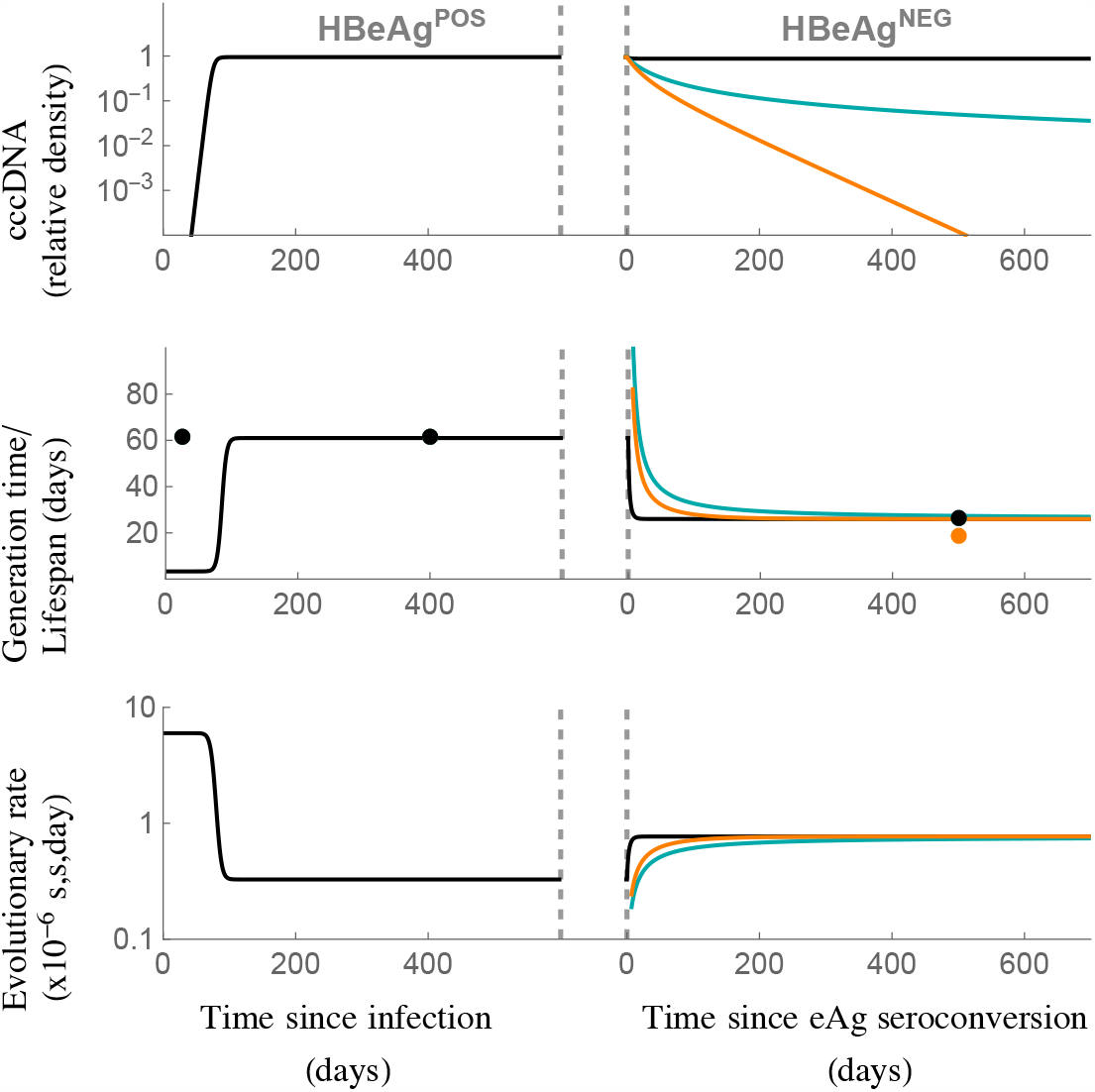
Population and evolutionary dynamics of cccDNA for the within-host model assuming all cccDNA survives mitosis (*q*=1). The model was parameterised assuming the generation time, *g*, during chronic HBeAg^POS^ infection is 61 days, and during chronic HBeAg^NEG^ infection is 26 days, in line with our predictions for cccDNA generation time *in vivo*. The top panel shows the cccDNA burden, where 1 represents the maximum possible burden. The middle panel shows the viral generation time (lines) and cccDNA lifespan during key stages of infection (dots, derived from Eqs 9 and 10). The bottom row shows the evolutionary rates. Black line: replicative capacity during HBeAg^NEG^ infection remains the same as during HBeAg-postive infection (*b*_*eAg+*_*= b*_*eAg-*_*=0*.*3* per day). Blue line: replicative capacity falls to *b*_*eAg-*_=0.038 per day during HBeAg^NEG^ infection, and *R*_*0*_*=1*. Orange line: replicative capacity falls to *b*_*eAg-*_*=0*.*038* per day and *R*_*0*_=0.7. See Table 1 for all other parameters.

**S2 Fig.**
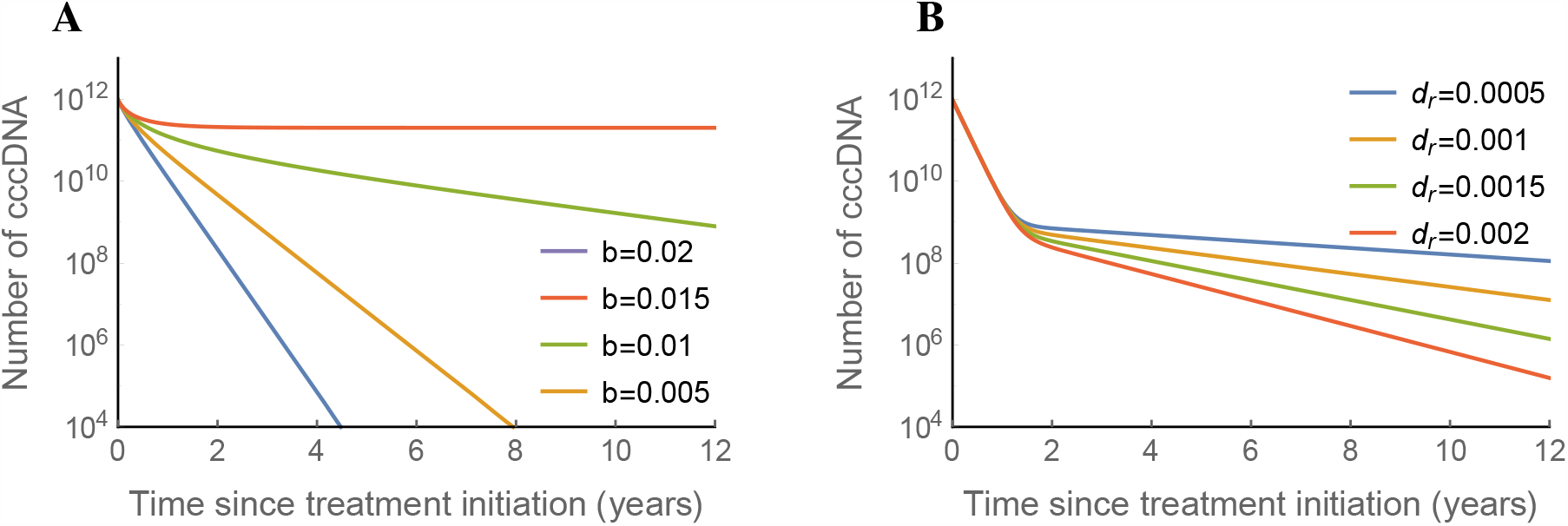
Effect of NA treatment predicted by the model assuming all cccDNA survives mitosis (*q*=1). A: cccDNA dynamics whilst on treatment, assuming some residual reproduction. For all cells *d*=0.002 per day, *δ*=0.014 per day, *c*=0, *q=1*. B: cccDNA dynamics on treatment, assuming no residual reproduction (*6*=0) but 0.1% cccDNA is long-lived, for different death rates of long-lived cells, *d*_*r*_ per day. For normal cells *d*=0.002, *δ*=0.014, *c*=0, *q*=1, and for long-lived cells *δr*=0, *C*_*r*_=0, *q*=0. The maximum number of cccDNA was assumed to be 10^12^, and all model runs were started at equilibrium in the absence of treatment (*b*=0.3).

